# Hemodynamic modeling of aspirin effects on BOLD responses at 7T

**DOI:** 10.1101/2020.01.30.20019729

**Authors:** Cao-Tri Do, Zina-Mary Manjaly, Jakob Heinzle, Dario Schöbi, Lars Kasper, Klaas P. Pruessmann, Klaas Enno Stephan, Stefan Frässle

## Abstract

Aspirin is considered a potential confound for functional magnetic resonance imaging (fMRI) studies. This is because aspirin affects the synthesis of prostaglandin, a vasoactive mediator centrally involved in neurovascular coupling, a process that underlies the blood oxygenated level dependent (BOLD) response. Aspirin-induced changes in BOLD signal are a potential confound for fMRI studies of patients (e.g. with cardiovascular conditions or stroke) who receive low-dose aspirin prophylactically and are compared to healthy controls that do not take aspirin. To examine the severity of this potential confound, we combined high field (7 Tesla) MRI during a simple hand movement task with a biophysically informed hemodynamic model. Comparing elderly volunteers with vs. without aspirin medication, we tested for putative effects of low-dose chronic aspirin on the BOLD response. Specifically, we fitted hemodynamic models to BOLD signal time courses from 14 regions of the human motor system and examined whether model parameter estimates were significantly altered by aspirin. While our analyses indicate that hemodynamics differed across regions, consistent with the known regional variability of the BOLD response, we neither found a significant main effect of aspirin (i.e., an average effect across brain regions) nor an expected drug×region interaction. While our sample size is not sufficiently large to rule out small-to-medium global effects of aspirin, we had adequate statistical power for detecting the expected interaction. Altogether, our analysis suggests that low-dose aspirin, as used for prophylactic purposes, does not strongly affect BOLD signals and may not represent a critical confound for fMRI studies.

## Introduction

Aspirin belongs to the group of non-steroidal anti-inflammatory drugs (NSAID) and is one of the most frequently used substances to reduce inflammation or pain (Vane, 1971; Vane and Botting, 2003). Due to its additional effect on thrombocyte aggregation it is commonly used in primary and secondary prevention of vascular disease (e.g. heart disease, stroke). It is known to inhibit cyclooxygenase (COX), an enzyme responsible for the production of prostaglandins (PG) through the conversion of arachidonic acid. The inhibition of COX results in a reduction of the synthesis of PG which, amongst other functions, serve to regulate contraction and dilation of vascular smooth muscle cells (Bolton, 1979). Notably, COX has different isoforms (COX-1 and COX-2) with differential and complex effects on vascular tone (Vanhoutte, 2009; Félétou, Huang and Vanhoutte, 2011), and the effect aspirin exhibits on COX is dose dependent (Warner, Nylander and Whatling, 2011). In low doses (less than 100mg/d) primarily COX 1 is inhibited. Intermediate to high doses (650mg − 8g/d) effectively inhibit both COX1 and COX2.

This has potential implications for fMRI since, in the brain, COX-dependent PG are involved in vasodilation in response to neural activity; for reviews, see (Lauritzen, 2005; Haydon and Carmignoto, 2006). This link between neural activity and vascular responses (neurovascular coupling) is an essential component in the generation of the blood oxygenated level dependent (BOLD) signal (Hillman, 2014; Huber *et al*., 2014). In brief, neural activity induces local functional hyperemia, i.e., an increase in regional cerebral blood flow (rCBF) in the vicinity that surpasses metabolic demand. This leads to an increase of oxygenated relative to deoxygenated hemoglobin which, in turn, changes the magnetic properties of blood (oxygenated hemoglobin is diamagnetic, while deoxygenated hemoglobin is paramagnetic) and thus the BOLD signal.

While the exact basis of neurovascular coupling is still subject to debate (Hillman, 2014), one potential mechanism concerns the increase of calcium in response to activation of glutamatergic receptors (Zirpel, Lachica and Rubel, 1995). This increase in calcium, in turn, leads to activation of phospholipase A_2_ (PLA_2_), with subsequent production of arachidonic acid that is converted to vasoactive prostaglandins by means of COX (Wang *et al*., 2006; Winship, Plaa and Murphy, 2007; Lind *et al*., 2013; Hillman, 2014). This chain of biochemical events suggests that an inhibition of COX by NSAID, like aspirin, and the ensuing reduction in vasodilatory prostaglandins could diminish blood flow and thus the BOLD signal.

While potential effects of aspirin on the BOLD response are relevant for any BOLD-fMRI study, this might be of particular concern for experiments with patients with cardiovascular conditions and/or stroke. These patients often require daily aspirin for secondary prophylaxis, yet are typically compared to healthy controls that are not matched for aspirin intake. This may induce a systematic bias when comparing the two groups (D’Esposito, Deouell and Gazzaley, 2003) and represents a general potential concern for comparing younger participants to elderly participants (who are more likely to receive prophylactic aspirin).

So far, studies examining the potential influence of COX inhibition on rCBF and BOLD signal have primarily focused on animals. For instance, both non-selective COX inhibition by indomethacin and selective inhibition of COX-2 by rofecoxib significantly reduced rCBF in rats (Bakalova, Matsuura and Kanno, 2002). Similarly, Stefanovic et al. found a significant decrease in rCBF as well as BOLD signal in rats by the preferential COX-2 inhibitor meloxicam (Stefanovic, Bosetti and Silva, 2006). Furthermore, decreases in resting rCBF were observed after administration of aspirin in rabbits (Bednar and Gross, 1999) and rats (Quintana *et al*., 1983).

While these animal studies fairly consistently demonstrate effects of COX inhibition (mainly via COX-2) on rCBF and BOLD signals, these experiments were performed with acute administration of NSAID, typically at high doses and mostly with drugs other than aspirin. In humans, a few studies of aspirin effects on CBF and/or BOLD have been performed (Markus, Vallance and Brown, 1994; Bruhn, Fransson and Frahm, 2001; Maihöfner *et al*., 2007; Kröger and May, 2014). Here, the picture is less clear. This is partially due to differences in methods and dosages, and partially because of small sample sizes and the use of pain stimuli, making it difficult to disentangle reduced nociception from decreased neurovascular coupling. Furthermore, to our knowledge, all existing studies administered aspirin acutely.

Hence, it presently remains unclear whether effects of chronic aspirin medication at typical prophylactic doses (e.g. 75-100 mg daily usage for stroke prevention) would substantially alter the BOLD signal in humans and thus represent a potential confound for fMRI studies.

To address this question, we used high field (7 Tesla) MRI to measure the BOLD signal during a simple hand movement task in healthy subjects who received aspirin for prophylactic purposes compared to an age-matched healthy control group without aspirin. The high magnetic field strength was chosen to exploit the high SNR afforded by 7T when testing for (potentially subtle) effects of aspirin. Similarly, we chose a simple motor task that evokes strong BOLD responses in multiple regions. To quantify aspirin effects on hemodynamics, we used a biophysically informed model. This hemodynamic model rests on an extension to the Balloon model (Buxton, Wong and Frank, 1998; Stephan *et al*., 2007) and enables inference on the temporal evolution of vasodilatory signal, blood flow, blood volume and deoxyhemoglobin contents from BOLD data.

To our knowledge, this study is novel in two ways: it is the first to examine chronic low-dose aspirin effects on BOLD responses, and it introduces the use of biophysically interpretable generative models to studying aspirin effects on BOLD responses. In addition to our model-based approach, we also consider conventional phenomenological parameters of the hemodynamic response function (HRF) (i.e., peak latency, peak amplitude, full width half maximum, that have frequently been used to characterize BOLD responses in the past (West *et al*., 2019).

## Methods

### Participants

30 age-matched volunteers (15 without aspirin intake [8 female, mean age: 60.5± 8.4 years], 15 with aspirin intake [6 female, mean age: 60.8 ± 10.6 years]) participated in the study. The aspirin group were selected based on their cardiovascular risk profile and took 100 mg aspirin per day for at least 2 weeks as a primary or secondary prevention for cardiovascular disease. Participants gave written informed consent to participate in the study. The study was conducted at the MR Center of the Institute for Biomedical Engineering, University of Zurich and ETH Zurich, at the University Hospital Zurich. The study conforms with the standards in the Declaration of Helsinki and was approved by the cantonal ethics committee Zurich under EK 09-2006 (ETH).

### Experimental design

Participants performed a simple motor paradigm, involving visually synchronized left (LH) and right hand (RH) fist closings. To make the task as simple as possible for our participants, the two hand movement conditions were separated into two scanning sessions. In each session, 14 blocks were presented, with 20 trials per block each trial, participants were instructed to fixate on a cross presented in the center of the screen, followed by a cue that indicated which hand to use in the upcoming trial. The Inter-Stimulus Interval (ISI) was set to 500 ms with a stimulus duration of 300 ms (yielding a trial length of 800 ms). Hence, each block lasted 16 seconds, and hand movement blocks were interleaved with a resting period of the same length where participants did not perform any hand movements. Stimuli were presented using Cogent 2000 (v1.33, http://www.vislab.ucl.ac.uk/cogent_2000.php).

### Data acquisition

The experiment was conducted on a 7 Tesla MR scanner (Phillips Achieva) with a 16-channel head coil. For each subject, we acquired 230 functional images per session (left and right hand movement) using a 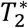-weighted echo planar imaging (EPI) sequence (TR = 2000s, TE = 25 ms, axial slices across the whole brain = 36, field of view (FOV) = 220 × 220 × 108 mm^3^, voxel size: 1.77 × 1.77 × 3 mm^3^, flip angle = 70 deg, SENSE factor 4). Additionally, an anatomical image was acquired by means of a T1-weighted inversion recovery turbo field echo (3D IR-TFE) sequence (TR = 7.7 ms TE = 3.5 ms, volume TR=4000 ms, inversion time 1200 ms, number of stacked slices = 150, voxel size: 0.9 × 0.9 × 0.9 mm^3^, FOV = 240 × 240 × 135 mm^3^, SENSE factor 2 in phase and 1.5 in slice direction). Simultaneous to the fMRI data acquisition, participants’ heart rate and respiration were recorded using a four electrode electrocardiogram (ECG) and a breathing belt, respectively.

### Data processing and analysis

The raw fMRI data were preprocessed using the open-software package SPM12 (v6685, Wellcome Trust Centre for Neuroimaging, London, UK, https://www.fil.ion.ucl.ac.uk/spm/) and MATLAB 2015a (Mathworks, Natick, MA, USA). Functional images were realigned, unwarped, coregistered to the participants’ individual anatomical image and normalized to the Montreal Neurological Institute (MNI) template space using the unified segmentation-normalization approach (Ashburner and Friston, 2005). The resulting images were then spatially smoothed using an isotropic Gaussian Kernel (FWHM: 8 mm).

The fMRI data were analyzed by means of a first level General Linear Model (GLM, (Friston *et al*., 1995)) with one task regressor, modeling the fist closings as events. This regressor was convolved with the canonical hemodynamic response function from SPM. Additionally, nuisance regressors were included to account for variance unrelated to the experimental manipulation. Specifically, six motion regressors (as obtained during the realignment) were included, as well as regressors accounting for cardiac and respiratory confounds obtained from the PhysIO Toolbox (Kasper *et al*., 2017), which is available as part of the open source TAPAS software (www.translationalneuromodeling.org/software) and implements the RETROICOR model (Glover, Li and Ress, 2000). Fourier expansions of third order for cardiac and fourth order for respiratory phases were used, as well as terms that account for the cardiac-respiratory interaction to model periodic effects of motion and field fluctuations. This yielded 18 physiological regressors which entered the fMRI first level GLM specification.

### Definition of regions of interest

Subsequently, a second level group analysis was specified to obtain the group maxima from an effects of interest F-contrast. We additionally computed brain activation maps using T-contrasts (LH > RH, RH > LH) to illustrate the well-established contralateral dominance of the motor network.

From the F-contrast, we identified 14 regions that showed significant whole-brain activation at the group level (*p* < 0.05, family-wise error (FWE) corrected at the peak level) associated with the task: primary motor cortex (M1), cerebellum (Cereb), thalamus (Thal), supplementary motor area (SMA), middle temporal visual area (hMT/V5), precentral gyrus (PcG) and insula, each in both hemispheres.

Voxel time series were extracted from left M1, Thal, SMA, V5, PcG, insula and right Cereb during the RH session, and from right M1, Thal, SMA, V5, PcG, insula and left Cereb during the LH session. For each subject and region, the BOLD signal time series were extracted as the principal eigenvariate of all voxels within a sphere of radius 4 mm (except for Thal where a radius of 2 mm was used to account for the small size of the area). The center coordinate of that sphere was identified for each subject as the individual nearest local maximum within a 12 mm sphere around the group activation peak. Notably, to avoid any overlap for the two SMAs (which are close to the longitudinal fissure), we constrained the region of interest to the respective hemisphere using an anatomical mask representing left and right hemisphere, respectively (WFUPICKATLAS toolbox, (Maldjian *et al*., 2003)). To quantify the effect of aspirin on the BOLD response, single-region hemodynamic models were then fitted to each of the extracted time series, separately.

### Computational Model

#### Hemodynamic Model

Our hemodynamic modeling approach is derived from the Dynamic Causal Modeling (DCM) framework for fMRI data (Friston, 2003). DCM is a generative model that distinguishes neuronal and hemodynamic states when fitted to measured BOLD signal time courses. The hemodynamic component (which is of particular interest in the present study) rests on the Balloon model (Buxton et al. 1998) and subsequent extensions (Friston al. 2000)(Stephan *et al*., 2007). The hemodynamic model itself can be separated into two components: First, neurovascular coupling describes the relative change in regional blood flow (rCBF) as a function of changes in neuronal activity (Friston *et al*., 2000):

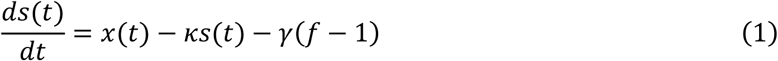

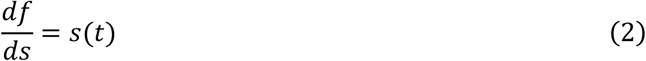

where *x* specifies neuronal population activity, *s* represent the vasodilatory signal, and *κ* and *γ* are rate constants of signal decay and feedback autoregulation, respectively. The variable *f* represents normalized (relative to rest) blood flow.

Second, changes in blood flow result in local changes in venous blood volume *v* and in deoxygenated hemoglobin content q (Buxton, Wong and Frank, 1998):

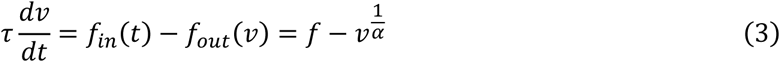

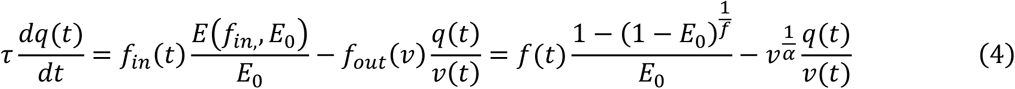

Here, *τ* is the mean transit time of blood which roughly corresponds to the ratio of resting blood volume *V*_0_ to resting blood blow *F*_0_. The dynamics of blood flow and deoxygenated hemoglobin content determine the measured BOLD signal. This is described by the BOLD signal output equation, a nonlinear function of the two biophysical quantities (Stephan *et al*., 2007):

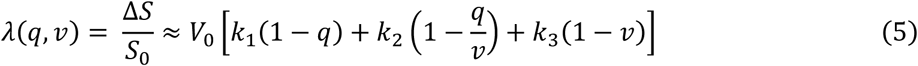

In this equation, *k*_1_,*k*_2_ and *k*_3_ are field strength dependent parameters and are given by *k*_1_ = 4.3*ϑ*_0_*E*_0_*TE, k*_2_= *εr*_0_*E*_0_*TE* and *k*_3_ = 1−*ε*. Here, *ϑ*_0_ is the frequency offset at the outer surface of magnetized vessels, *E*_0_ the oxygen extraction fraction at rest, *TE* the echo time, *r*_0_ the intravascular relaxation rate of oxygen saturation and *ε* represents the ratio between intravascular and extravascular MR signal (for more information, see Appendix A in (Heinzle *et al*., 2016) or (Stephan *et al*., 2007)). As mentioned above, in DCM for fMRI, the hemodynamic model is coupled to the neuronal model which describes effective (directed) connectivity among neuronal populations. In the present study, we were only interested in the hemodynamic properties (e.g., *τ, κ*) of multiple regions, not their connectivity. This, however, still requires modeling how neuronal events trigger vascular processes. One option would be to follow the approach of voxel-wise general linear models (GLM) and feed simple representations of neuronal activity (events or blocks) into the neurovascular coupling equation (Eq. 1). This approach was chosen in earlier work (Friston et al. 2000). Here, we extended this approach and considered a minimal neuronal model that captures some basic response properties of neuronal populations such as the self-dampening nature of induced transients (compare (Miller *et al*., 2001)). Effectively, we fitted single-region DCMs to BOLD signal from each region separately (**Fig. 1**) but omitted bilinear and non-linear terms (of how inter-regional connections are modulated) from the neuronal state equations. This yielded the following simplified neuronal model for a single region:

**Fig 1:**
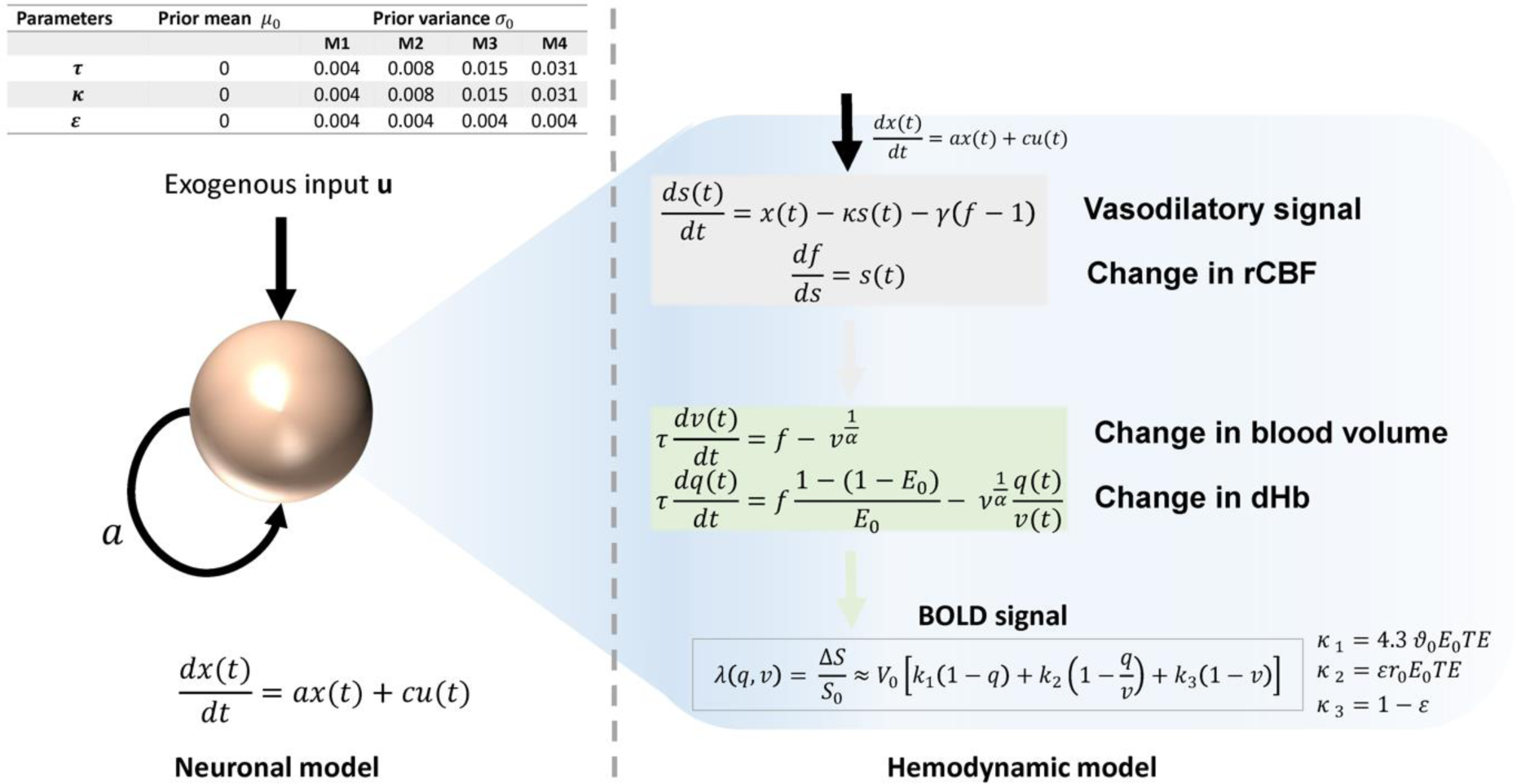
Left upper panel: Summary of the priors on the free parameters of the hemodynamic model. These parameters are specified in log space. *Left bottom*: The neuronal state equation for the single region DCM; *Right bottom*: State equations of the hemodynamic model that can be partitioned into two components: First, neural activity generates a vasodilatory signal and causes resting cerebral blood flow to change. Second, changes in blood volume and deoxygenated hemoglobin into a nonlinear output equation of the predicted BOLD signal.

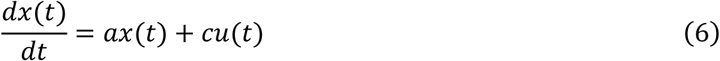

Here, *a* represents the rate constant of neuronal self-dampening (equivalent to an inhibitory “self-connection”) in a single region. *c* represents a weight factor for the driving input (e.g. sensory stimuli). Furthermore, to adequately account for different acquisition timings between slices, sampling times were computed for each of the regions of interest and taken into account as delays in the observation equation; see (Kiebel *et al*., 2007).

### Settings of DCM

The settings of the hemodynamic and neuronal model for the single-region DCM were based on the default settings in SPM 12 (v6560), with several notable exceptions to make the model suitable for our research question: First, as the focus of standard DCM is on the neuronal (i.e., effective connectivity) parameters, the priors on the hemodynamic parameters are relatively narrow. In contrast, the present study explicitly focuses on the hemodynamic parameters and thus requires less informed priors. To account for uncertainty about prior variance, we used several values of prior variances for each model inversion and subsequently marginalized over these prior variances (Bayesian Model Averaging (BMA; see below). Specifically, we scaled the default prior variance of the transit time *τ* and decay parameter *κ* of the hemodynamic model by a multiplicative factor (i.e., [1, 2, 4, 8]).

Second, as the parameters *k*_1_, *k*_2_ and *k*_3_ in the BOLD signal output equation (see Eq., 5) depend on the magnetic field strength, we adjusted these parameters to the values reported for 7T (Heinzle *et al*., 2016). Third, the parameter *ε* (see **Fig. 1**; right bottom panel) is part of *k*_2_and *k*_3_ and thus not a direct component of the model describing regional hemodynamics. Hence, the prior mean and variance of *ε* were chosen to be the same in all models (see **Fig. 1**; top left panel). Finally, the intrinsic self-connection (i.e., parameter *a*) was fixed (by setting the prior variance to 0) to a value of −0.5*exp(3). This value was chosen in order to obtain fast neural transients and thus limit the contribution of the neuronal level.

### Variational Bayesian Inference

In order to infer the hidden states and parameters, model inversion was performed using variational Bayes under the Laplace assumption (VBL)(Friston *et al*., 2007) as implemented in SPM12. In order to (at least partly) overcome the well-known local extrema problem of VB schemes, a multi-start approach was used by spanning a search grid of starting values. The values were chosen relative to the default prior variance; specifically, starting values were chosen as multiples of the standard deviation for the transit *τ*[-sqrt(8), − sqrt(4), −sqrt(2), −1, 0, 1, sqrt(2), sqrt(4), sqrt(8)] and the rate constant *κ*[-sqrt(8), −sqrt(4), −sqrt(2), −1, 0, 1, sqrt(2), sqrt(4), sqrt(8)]. The starting values of the driving input C were set either to 0 or 1. This choice reflects the expected positive input due to selection of positively activated regions in the GLMs. For each model, this yielded 162 different combinations of starting values for each model. Altogether, this resulted in 162 starting value combinations * 4 models per region* 14 regions = 9072 model inversions per subject. For a given model, of all starting values, the inversion yielding the highest model evidence was then chosen. The respective estimates of parameters and model evidence were then used for further statistical analysis.

#### Bayesian Model Averaging

To deal with model uncertainty and marginalize over prior variances, Bayesian Model Averaging (BMA, (Penny *et al*., 2010)) was performed over the four models that differed in the prior variance for the transit and decay parameters, as described above. In order to obtain estimates of the parameters across models, for each subject and region, a model-independent posterior estimate was obtained by marginalizing over models weighted by the posterior probability of each model:

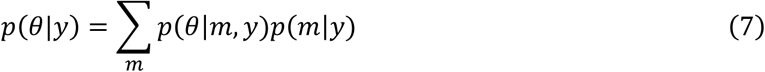

### Phenomenological parameters of the HRF

In addition to BMA estimates of parameters from our hemodynamic model, we also considered a model-independent approach. For this purpose, we used three more classical features of estimated hemodynamic response functions (HRF) (**Fig. 2**): the peak latency (P_L_), the peak amplitude (P_A_) and the full width at half maximum of the HRF (FWHM). These features of the HRF have previously been used to characterize population differences in hemodynamic responses, for example, in the context of healthy aging (West *et al*., 2019). These phenomenological parameters were computed by using the BMA parameter estimates of decay and transit parameters for reconstructing region-specific HRFs from the first order Volterra kernel using the function spm_kernels.m (v6937).

**Fig 2:**
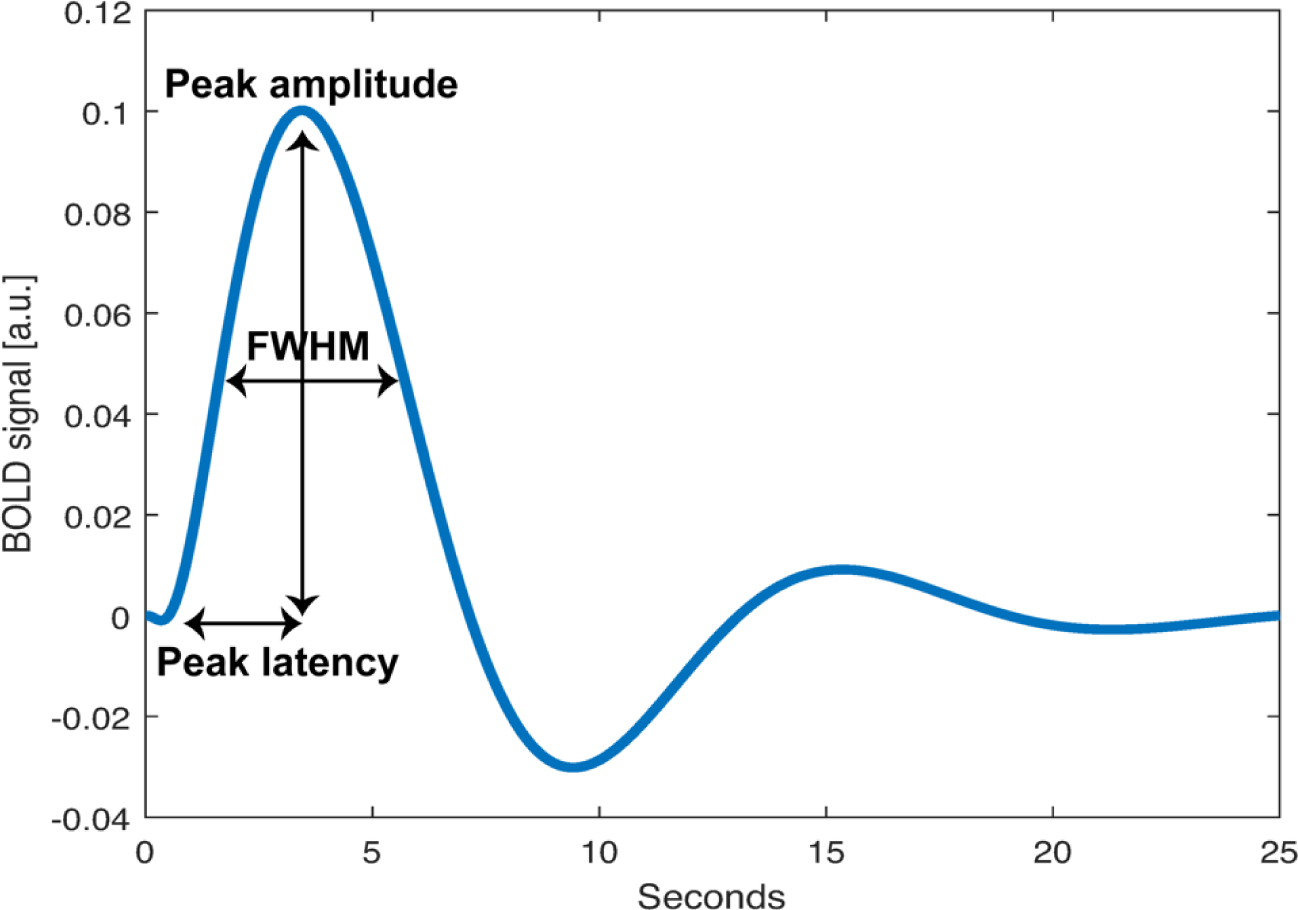
Sample of Hemodynamic response function (HRF) for one subject and region (Right Primary Motor Cortex, M1) reconstructed from the decay and transit parameters of the hemodynamic model computed from the first order Volterra Kernel. Three phenomenological parameters were included in the analysis: timing of the peak amplitude (time2peak), full width at half maximum of the HRF (FWHM), magnitude of the peak amplitude (Peak).

### Statistical comparison

To explore putative effects of aspirin on the HRF, the BMA estimates of the biophysical parameters of the Balloon model (i.e., rate constant *κ* and transit *τ*) as well as the phenomenological HRF parameters were then subjected to statistical tests. Specifically, for each parameter estimates, a mixed-effects repeated-measures ANOVA design was used, including a within-subject factor (“region”) and a between-subject factor (“drug”), running under R Studio (v. 1.2.1335). Prior to the statistical analysis, Mauchly’s test was used to check the validity of the sphericity assumptions (Mauchly, 1940). In case these were violated, the degrees of freedom were corrected using Greenhouse Geisser estimates of sphericity (Geisser and Greenhouse, 1959).

## Results

### BOLD activity during unilateral hand movements

Visually synchronized unilateral hand movements engaged a widespread network of cortical and subcortical regions, mainly lateralized towards the contralateral hemisphere (**Fig. 3**; *p* < 0.05, FWE-corrected at the peak level for multiple comparisons). In this, five participants were excluded from the analysis because they did not perform the task correctly (i.e., they closed the fist continuously instead of alternating between opening and closing hand movements) or did not show any motor activity (i.e., during the fist clench condition only visual areas were activated). Overall, activation was most pronounced in the following regions: primary motor cortex (M1), cerebellum (Cereb), thalamus (Thal), supplementary motor area (SMA), middle temporal visual area (hMT/V5), precentral gyrus (PcG) and insula (see Table 1). These regions were chosen for subsequent generative modeling of the hemodynamic responses.

**Fig 3:**
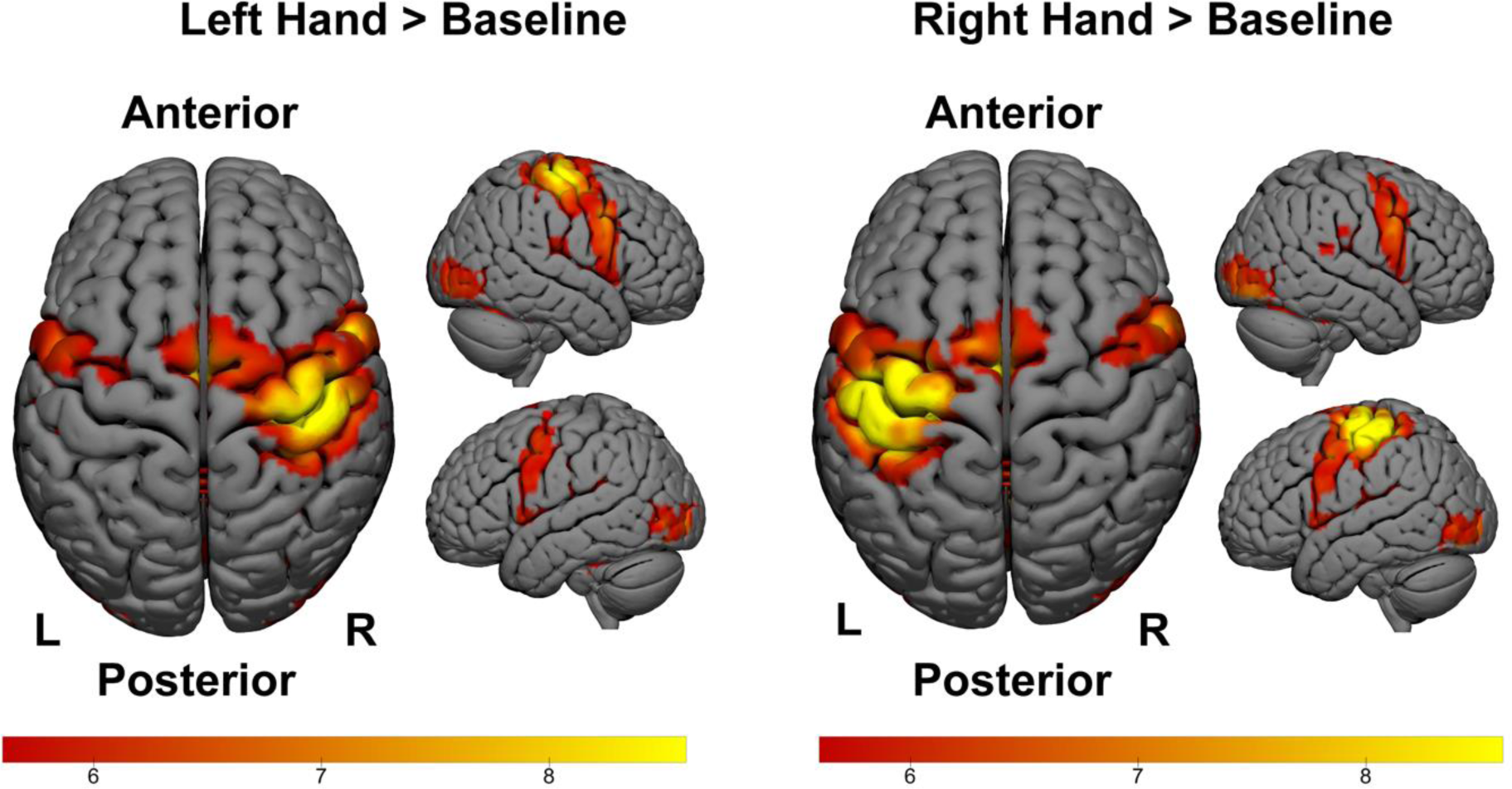
Activation maps of the two T-contrasts (LH > Baseline and RH > Baseline) obtained from a second level group analysis; Several regions were identified that are significantly activated (whole brain correction p<0.05, family-wise error (FWE) corrected at the group level) during the hand movement tasks: primary motor cortex (M1), Cerebellum (Cereb), Thalamus (Thal), Supplementary Motor Area (SMA), middle temporal visual area (hMT/V5), Precentral Gyrus (PcG) and Insula, each in both hemispheres.

**Table 1:**
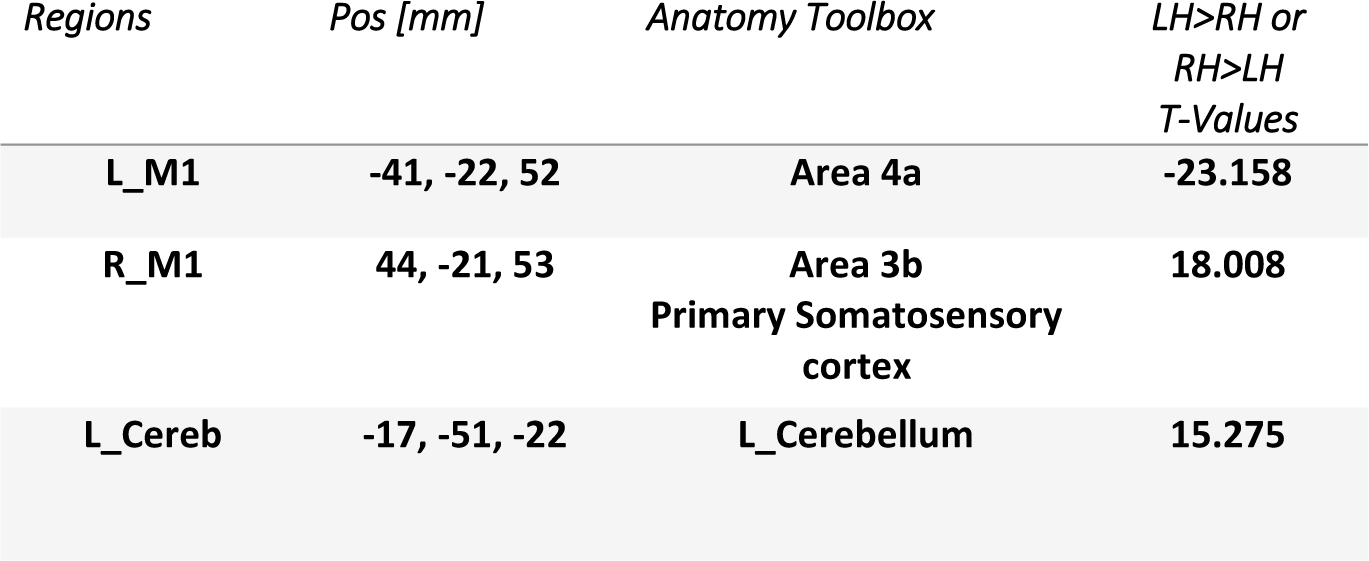
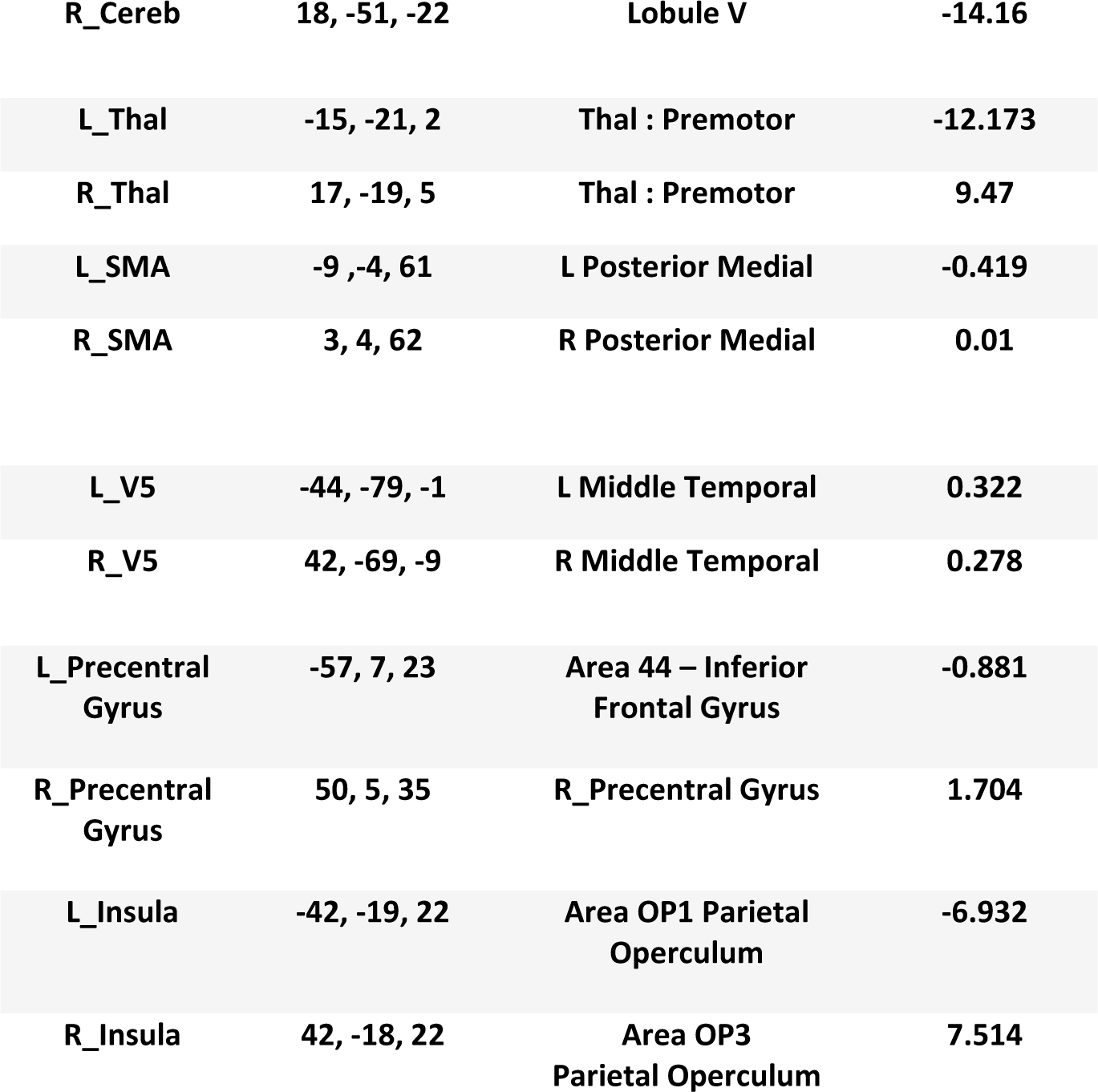
Regions of interest which showed significant BOLD activation during visually synchronized unilateral hand movements and were therefore subjected to subsequent DCM analyses. The labels for the brain regions were taken from the SPM Anatomy Toolbox (v2.2b). Here, the positive T-Values reflect left hand favored whereas negative T-Values are right hand favored.

### Hemodynamic modelling through single-region DCMs

Single-region DCMs were then fitted to the BOLD signal time series extracted from the regions of interest mentioned above in order to infer hemodynamic parameters (see Methods, **Fig. 4**). The means of the BMA posterior densities for the mean transit time of blood and the rate constant of the signal decay (Stephan *et al*., 2007) as well as the phenomenological HRF parameters (peak latency, peak amplitude, and FWHM), were examined for significant differences (**Fig. 5-6**) between the two groups (aspirin vs no-aspirin) using mixed-effects repeated-measures ANOVAs (see Table 2).

**Fig 4:**
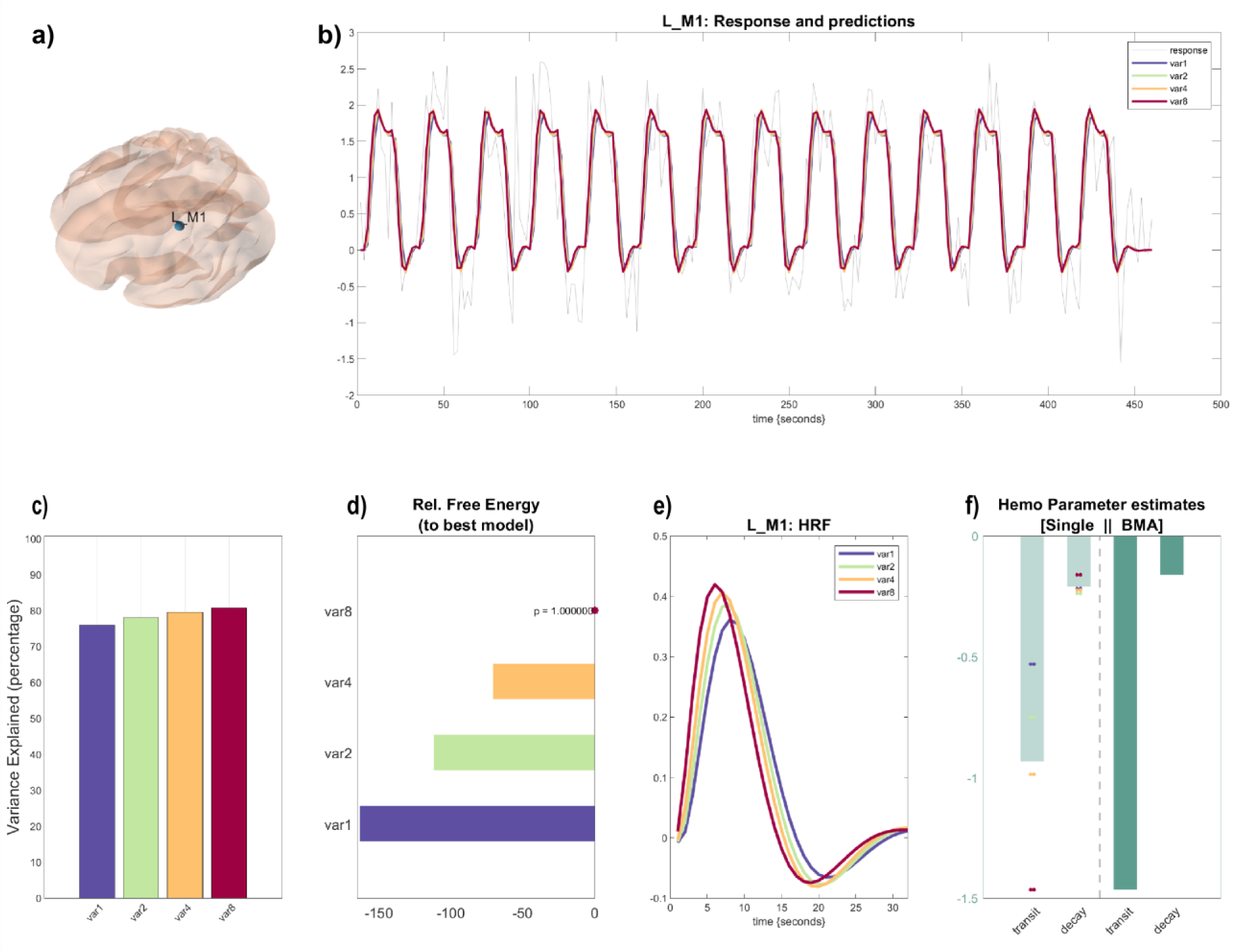
Example of a DCM inversion summary; For each subject we inverted 4 models that differed in their prior variances for the decay and transit parameters; *a*) Peak location of the cortical regions at which the voxel time series was extracted; *b*) Overview of model fits for the different DCMs; Gray: BOLD time series data obtained from the voxel time series extraction, In Color: Model fits for the different models; Here the predicted responses are overlapping; *c*) Variance explained for each model; *d*) Free energy relative to the best model, Star depicts best model (in terms of free energy); The pvalue describes the posterior probability of the winning model; e) Hemodynamic response function (HRF) reconstructed from first order Volterra kernel using the decay and transit parameter obtained from model inversion; *f*) Results from the Bayesian Model Averaging (BMA) for the decay and transit parameter; Light green bars represent mean estimates over single parameter inversions across the variances; Dark green bars are the estimates obtained after performing BMA.

**Fig 5:**
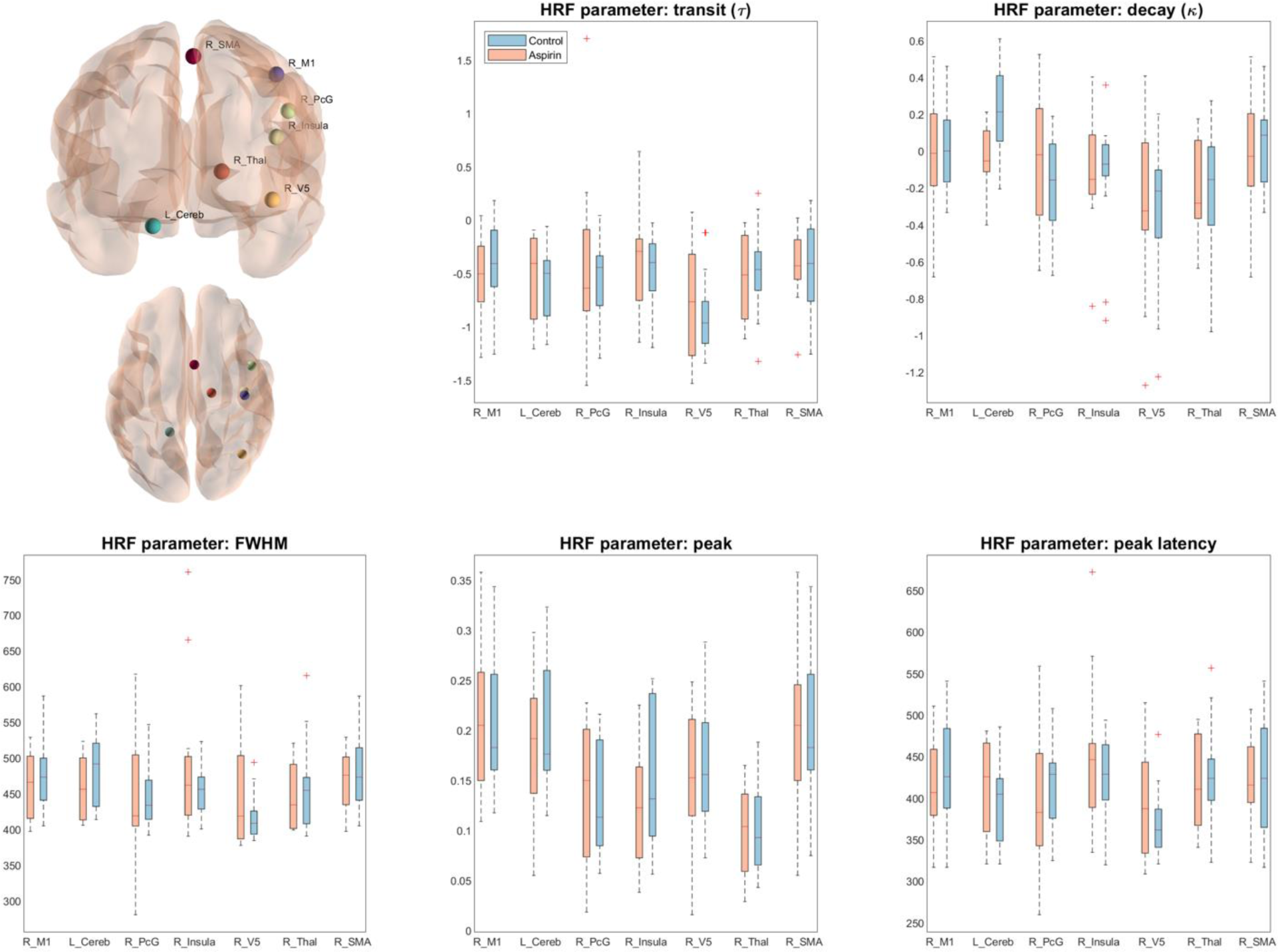
Posterior estimates from DCM inversion as well as the phenomenological parameters for the regions of interest that showed a significant positive T-Value (obtained from the group level analysis) in the left hand movement. Regions illustrate the group maxima obtained from the second level group statistics and solely meant for visualization.

**Fig 6:**
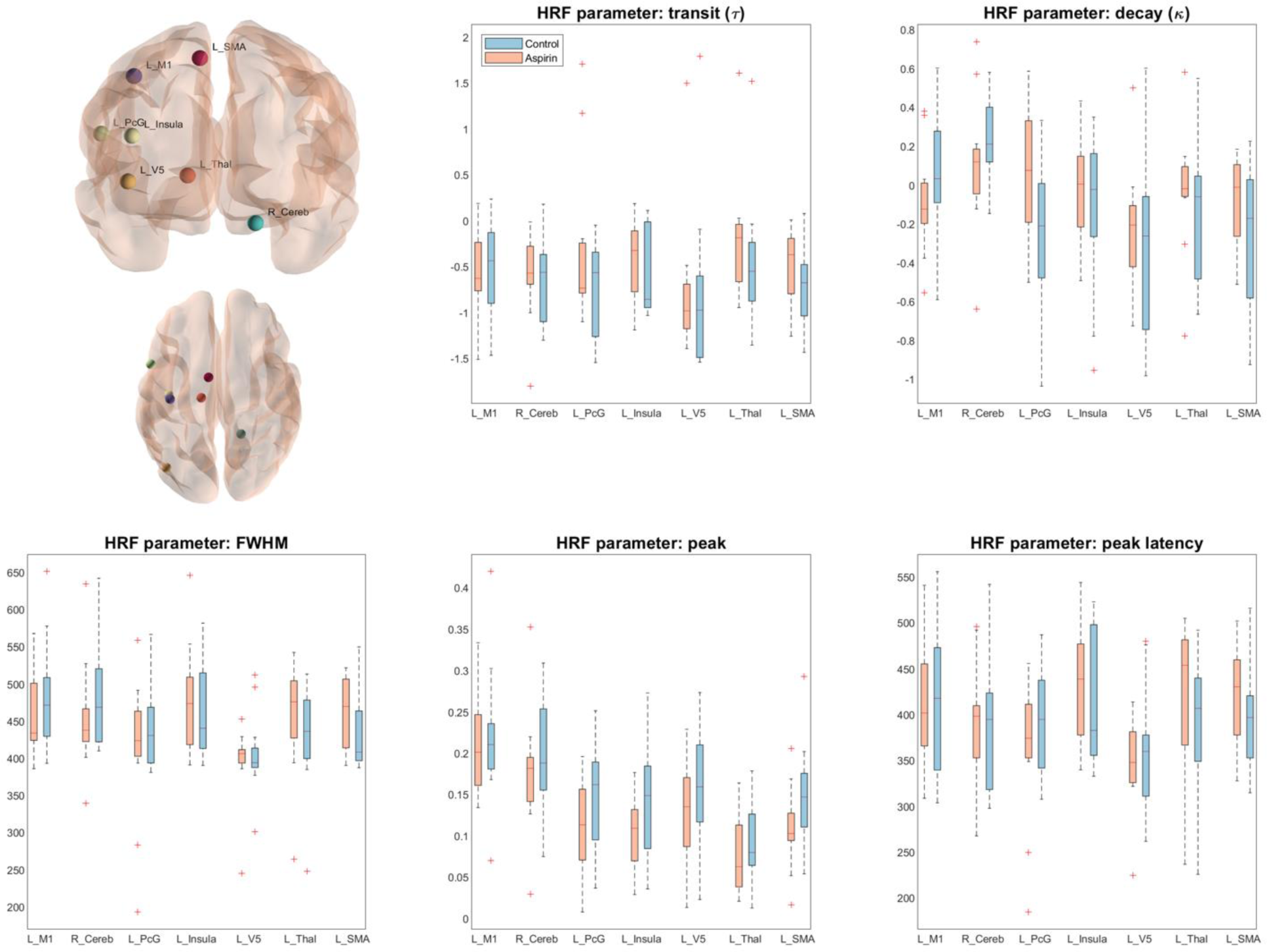
Posterior estimates from DCM inversion as well as the phenomenological parameters for the regions of interest that showed a significant positive T-Value (obtained from the group level analysis) in the right hand movement. Regions illustrate the group maxima obtained from the second level group statistics and solely meant for visualization.

**Table 2:** Results from the mixed-effects repeated-measures ANOVA for the different parameters of interest: decay, transit, peak latency, FWHM, and peak amplitude. The effect size are given in *η*^2^following the convention in (Bakeman, 2005) as well as in Cohen’s f. All values for the significance level are Greenhouse Geisser (GG) corrected.

| *Parameter* | *Effect* | *dF* | *F-statistic* | *P-Values* | *Effect size*<br>$\eta^2$ | *Effect size*<br>Cohen's $f$ |
| --- | --- | --- | --- | --- | --- | --- |
| *Decay* | Main effect: Region | 5.0, 115.05 | 6.245 | 0.000 (GG corrected) | 0.142 | 0.445 |
|  | Main effect: Drug | 1,23 | 0.126 | 0.726 (GG corrected) | 0.002 | 0.045 |
| | Interaction: Region × Drug | 5.0, 115.05 | 2.028 | 0.08 (GG corrected) | 0.051 | 0.225 |
| *Transit* | Main effect: Region | 4.52, 104.04 | 1.885 | 0.110 (GG corrected) | 0.056 | 0.313 |
|  | Main effect: Drug | 1,23 | 0.84 | 0.369 (GG corrected) | 0.01 | 0.009 |
| | Interaction: Region × Drug | 4.52, 104.04 | 0.609 | 0.457 (GG corrected) | 0.019 | 0.16 |
| *Peak latency* | Main effect: Region | 5.03, 115.61 | 4.178 | 0.002 (GG corrected) | 0.095 | 0.376 |
|  | Main effect: Drug | 1,23 | 0.058 | 0.812 (GG corrected) | 0.001 | 0.063 |
| | Interaction: Region × Drug | 5.03, 115.61 | 0.931 | 0.464 (GG corrected) | 0.023 | 0.11 |
| *FWHM* | Main effect: Region | 5.57, 128.09 | 4.265 | 0.000 (GG corrected) | 1.005 | 0.376 |
|  | Main effect: Drug | 1.23 | 0.005 | 0.945 (GG corrected) | 0.000 | 0.045 |
| | Interaction: Region × Drug | 5.57, 128.09 | 1.249 | 0.288 (GG corrected) | 0.032 | 0.157 |
| *Peak Ampl.* | Main effect: Region | 5.17, 118.89 | 17.576 | 0.000 (GG corrected) | 0.308 | 0.645 |
|  | Main effect: Drug | 1.23 | 1.024 | 0.322 (GG corrected) | 0.018 | 0.123 |
| | Interaction: Region × Drug | 5.17, 118.89 | 0.58 | 0.72 (GG corrected) | 0.014 | 0.128 |

For the decay parameter, we found a significant main effect of region (F(5.0,115.05) = 6.245, p<0.001) suggesting that the BOLD response differed considerably across regions, irrespective of aspirin intake. The the main effect of drug (F(1,23) = 0.126, p=0.726) was not found to be significant as well as the drug×region interaction (F(5.0,115.05) = 2.028, p=0.08). For the transit parameter, there was no significant main effect of region (F(4.52,104.04) = 1.885, p=0.11) as well as no significant main effect of drug (F(1,23) = 0.84, p=0.369) nor drug×region interaction (F(4.52,104.04) = 0.609, p=0.457).

For the phenomenological parameters, the results were similar. For the peak latency P_L_, there was a significant main effect of region (F(5.03,115.61) = 4.178, p=0.002), but no significant main effect of drug (F(1,23) = 0.058, p=0.812) or drug×region interaction (F(5.03,115.61) = 0.931, p=0.464). Similarly, for FWHM, the main effect of region was significant (F(5.57,128.09) = 4.265, p<0.001), but neither the main effect of drug (F(1,23) = 0.005, p=0.945) nor the drug×region interaction (F(5.57,128.09) = 1.249, p=0.288). Finally, for the peak amplitude P_A_, there was, once again, a significant main effect of region (F(5.17,118.89) = 17.576, p<0.001), but no significant main effect of drug (F(1,23) = 1.024, p=0.322) and no drug×region interaction (F(5.17,118.89) = 0.58, p=0.72).

As is generally the case for frequent statistics, the failure to reject a null hypothesis does not mean that we can conclude the absence of an effect. However, negative findings can be more easily interpreted in the light of a *s*tatistical power analysis. In our case, power analysis (using G Power v.3.1.9.2, (Erdfelder *et al*., 2009)) across different effect sizes (**Fig. 7**) indicated high power (i.e. on the order of 80%) for the main effect of region and the interaction even for small effect sizes (f = 0.15, following the effect size convention by Cohen (Cohen, 2013)). By contrast, for the main effect of drug, our experimental design yielded sufficient statistical power only for relatively large effect sizes (f ≥ 0.35). In other words, our analysis had high power to detect regional differences in aspirin effects on BOLD, but was less sensitive to an average effect of aspirin across regions.

**Fig 7:**
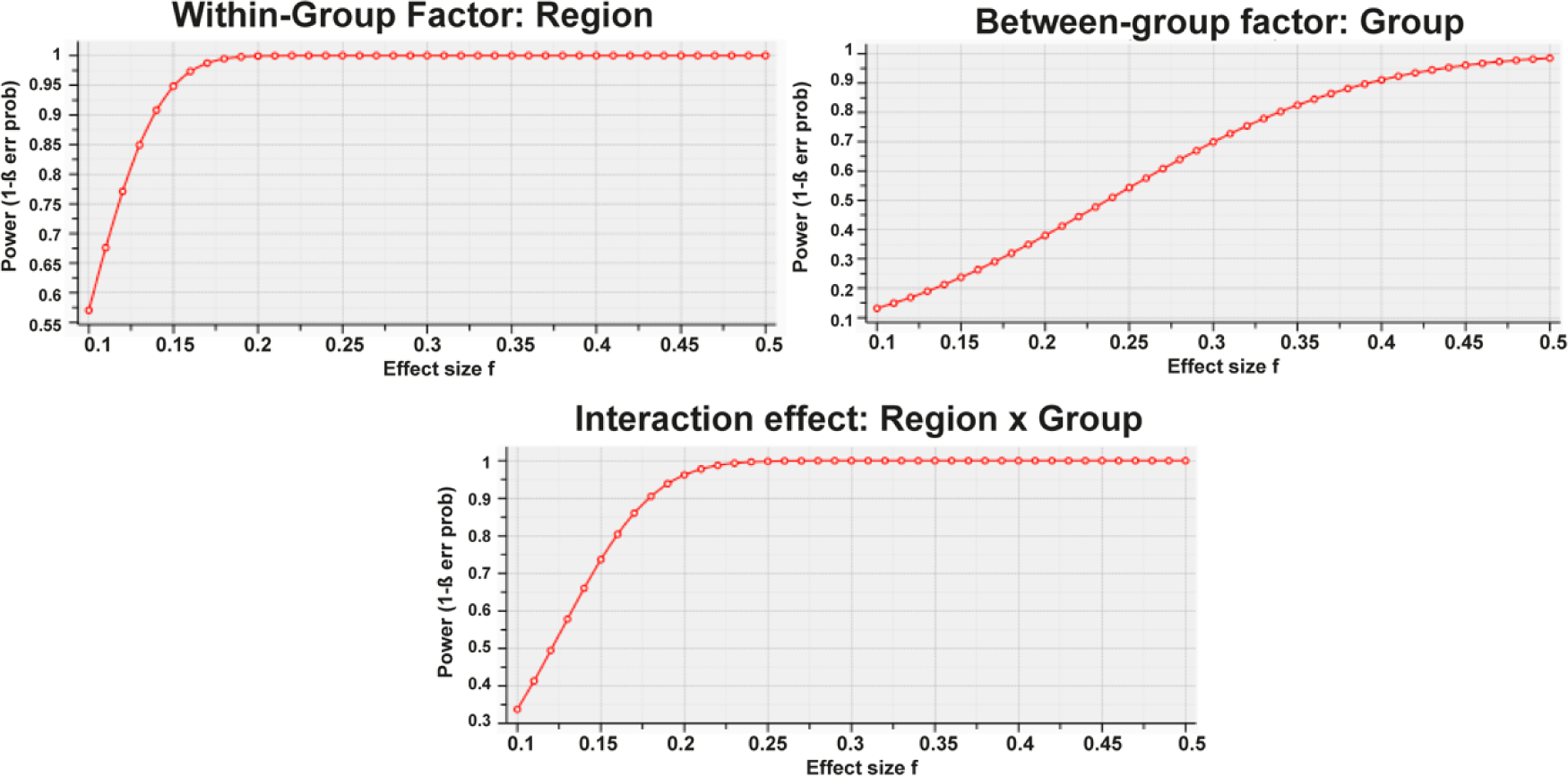
Statistical power analysis for the within-group, between-group factors and their interaction for different effect sizes

## Discussion

In this study, we investigated the effect of aspirin on hemodynamic responses in humans. Our study is novel in two ways: it examines chronic low-dose aspirin effects on BOLD responses, and it introduces a novel analysis approach, i.e., generative models of regional BOLD signals with a biophysical interpretation.

Our study followed a case-controlled design, contrasting elderly volunteers with versus without aspirin medication for prophylactic purposes. We attempted to maximize the signal-to-noise ratio of our BOLD measurements by (i) using a simple hand movement paradigm known to elicit strong activations in multiple brain regions, and by (ii) acquiring data at high (7T) field strength. Statistical analyses based on estimates from our biophysically informed hemodynamic model and on conventional phenomenological indices of the HRF, respectively, came to equivalent conclusions: while we observed that hemodynamic parameters (except for the transit time) differed considerably across brain regions (a main effect of region), we found no significant drug×region interaction and no significant main effect of drug (i.e., aspirin vs. no aspirin).

The observed main effect of region on the hemodynamic parameter estimates (as well as on the phenomenological HRF indices) is consistent with previous work indicating considerable variability of the hemodynamic response across regions and individuals (Aguirre, Zarahn and D’Esposito, 1998; Handwerker, Ollinger and D’Esposito, 2004; Handwerker *et al*., 2012). Specifically, hemodynamics has been shown to vary up to a second across different brain regions, e.g., from visual to frontal cortex (Buckner *et al*., 1996). Hemodynamic variability was found to be even more pronounced across different subjects (Aguirre, Zarahn and D’Esposito, 1998).

Our main question – the putative influence of aspirin on BOLD responses – has two facets. First, the main effect of drug: is there a “global” effect of aspirin on hemodynamics, i.e., an average effect across all regions tested? Second, the drug×region interaction: does the putative effect of aspirin on BOLD responses differ across regions? Concerning the latter, it is worth pointing out that, from a neurobiological perspective, regional variability in aspirin effects on neurovascular coupling would seem likely. This is because the constitutive (i.e., physiological, unrelated to inflammation) expression of both COX-1 and COX-2 varies across different brain regions in humans (Yasojima *et al*., 1999; Yermakova *et al*., 1999) and animals (Tsubokura *et al*., 1991; De Vries *et al*., 2003; Oláh *et al*., 2012). Notably, our “repeated measures” design (with multiple regional BOLD measures per subject) afforded high statistical power for testing the drug×region interaction, even for small effect sizes (**Fig. 7**). Our finding that the interaction was non-significant thus renders it unlikely that low-dose aspirin could have a sizeable influence on BOLD signals, in a manner that would be in accordance with neurobiological constraints.

However, from our results, we cannot exclude the possibility of a global effect of low-dose aspirin on hemodynamics: While the non-significant main effect of drug in combination with the results from our statistical power analysis makes a strong global effect of aspirin (that would have relevance for fMRI studies with patients) unlikely, our current study lacks the sensitivity to detect global influences of aspirin that are of medium or small effect size (**Fig. 7**).

Apart from the above-mentioned lack in sensitivity due to small sample sizes and low dosage, there are other potential reasons why we did not observe a significant effect of aspirin. First, our understanding of the mechanisms by which aspirin might influence hemodynamics is likely to be incomplete. This is because while various candidate mechanisms of neurovascular coupling (i.e., changes in hemodynamics generated by neural stimulation) have been proposed, a clear consensus is still missing. Several studies and reviews have highlighted the complexity of the relationship between neurovascular coupling and BOLD response (Hillman, 2014; Wright, Wise and Harris, 2018), as well as the plethora of neurovascular agents involved (for an overview, see (Riera and Sumiyoshi, 2010). Recently, it has been suggested that endothelial cells also play an important role in mediating vasodilatory activity through their vasoactive agents (Chen *et al*., 2014; Hillman, 2014). In summary, both fast (Wölfle *et al*., 2011) and slow (Tallini *et al*., 2007) components of neuronally induced vasodilation have been proposed, describing a substantial variety of biophysical and biochemical processes caused by the initial neuronal impulse.

As a consequence, it is presently difficult to formulate a hemodynamic model that captures all possible effects. The generative model of hemodynamic responses used in our study represents a principled and widely used model, but does not account for all facets of neurovascular processes, such as transient uncoupling between blood flow and blood volume (Mandeville *et al*., 1998; Chen and Pike, 2009; Kim and Ogawa, 2012; Huber *et al*., 2014) or the differential role of excitatory and inhibitory neurons. Recent developments have started to address these limitations. In particular, Havlicek et al. (Havlicek *et al*., 2015) introduced a variant of the hemodynamic model in DCM that aims at a more faithful representation of physiological processes. It is also worth pointing out, however, that the exact formulation of the hemodynamic model is unlikely to have played a decisive role for our results. This is because our non-model-based analysis, using conventional descriptive indices of the shape of the HRF, gave perfectly consistent results and also failed to reveal a significant main effect of drug or an interaction.

Another potential limitation of the present study is the advanced age of our subjects. It is known that the cerebrovascular system changes over the lifespan, resulting in changes in the structural vasculature. For instance, arteriosclerotic changes cause an alteration of blood vessel elasticity (Farkas and Luiten, 2001) and a decrease in capillary density (Meier-Ruge *et al*., 1980; Brown and Thore, 2011) resulting in changes of neurovascular coupling. For example, the signal-to-noise ratio of the BOLD signal during a simple sensorimotor task was found to be significantly decreased in elderly subjects compared to a younger control group (D’Esposito *et al*., 1999). These findings, in line with other studies (Hesselmann *et al*., 2001; Huettel, Singerman and McCarthy, 2001), suggest that neural activity and BOLD signal change notably with age. Furthermore, between- and within-subject variability of BOLD responses is increased in the older population, aggravating the interpretation of fMRI studies in this population (Kannurpatti *et al*., 2010; Baum and Beauchamp, 2014).

These limitations notwithstanding, the present study illustrates how high-field (7T) fMRI and biophysically informed modeling can be used to study pharmacological effects on the BOLD signal. While relevant for neuroimaging studies in general, for example with regard to formulation of exclusion criteria (compare D’Esposito et al. 2003), the question of whether aspirin effects BOLD is of particular importance for studies with patients who receive prolonged low-dose aspirin medication for reasons of primary or secondary prophylaxis. Altogether, our results suggest that strong effects of low-dose aspirin on BOLD signals are not likely. Given the limited statistical sensitivity of our analyses for certain (but not all) tests, our current results will require replication in future studies using larger samples. This should be feasible, given the emergence of large-scale databases combining both fMRI and health data from the general population (e.g., UK Biobank, (Sudlow *et al*., 2015)). The model-based approach presented in this study may serve as a useful tool for clarifying the practical impact of aspirin on fMRI studies.

## Data Availability

Upon acceptance of this paper, the code used for the analysis as well as the BOLD signal time series data will be made publicly available. Furthermore, the code has been cross-checked internally for reproducibility. All analysis streams were performed on the Euler cluster at ETH Zurich. More information on the computational power can be found at https://scicomp.ethz.ch/wiki/Euler.

## Acknowledgements

This work was supported by the René and Susanne Braginsky Foundation (KES) and the University of Zurich (KES) as well as by the ETH Zurich Postdoctoral Fellowship Program (SF), the Marie Curie Actions for People COFUND Program (SF), and the University of Zurich Forschungskredit Postdoc (SF).

## Conflict of Interest

Authors report no conflict of interest.

## Author contributions

CTD and ZMM contributed equally to the project. ZMM and KES designed the study. ZMM and LK conducted the experiment. CTD set up the data analysis pipeline including data preprocessing, modeling and statistical analysis. JH and SF provided assistance for the data analysis. SF performed the code review for the data analysis pipeline. DS provided input to the manuscript as well as to the optimization of the modeling approach. CTD, JH, SF and KES critically discussed and interpreted the results. CTD, JH, KES, and SF wrote the paper. ZMM, LK, DS, and KPP gave additional feedback on the manuscript.

## Notes

### Competing Interest Statement

The authors have declared no competing interest.

## References

1 Aguirre, G. K., Zarahn, E. and D’Esposito, M. (1998) ‘The variability of human, BOLD hemodynamic responses’, NeuroImage, 8(4), pp. 360–369. doi: 10.1006/nimg.1998.0369.

2 Ashburner, J. and Friston, K. J. (2005) ‘Unified segmentation’, NeuroImage, 26(3), pp. 839–851. doi: 10.1016/j.neuroimage.2005.02.018.

3 Bakalova, R., Matsuura, T. and Kanno, I. (2002) ‘The cyclooxygenase inhibitors indomethacin and Rofecoxib reduce regional cerebral blood flow evoked by somatosensory stimulation in rats’, Exp Biol Med (Maywood), 227(7), pp. 465–473. Available at: http://www.ncbi.nlm.nih.gov/entrez/query.fcgi?cmd=Retrieve&db=PubMed&dopt=Citation&list_uids=12094010.

4 Bakeman, R. (2005) ‘Recommended Effect Size Statistic’, Behavior Research Methods, 37(3), pp. 379–384. doi: 10.3758/BF03192707.

5 Baum, S. H. and Beauchamp, M. S. (2014) ‘Greater BOLD variability in older compared with younger adults during audiovisual speech perception’, PLoS ONE, 9(10). doi: 10.1371/journal.pone.0111121.

6 Bednar, M. M. and Gross, C. E. (1999) ‘Aspirin reduces experimental cerebral blood flow in vivo’, Neurological Research, 21(5), pp. 488–490. doi: 10.1080/01616412.1999.11740963.

7 Bolton, T. B. (1979) ‘Mechanisms of action of transmitters and other substances on smooth muscle’, Physiological Reviews, 59(3), pp. 606–718. doi: 10.1152/physrev.1979.59.3.606.

8 Brown, W. R. and Thore, C. R. (2011) ‘Review: Cerebral microvascular pathology in ageing and neurodegeneration’, Neuropathology and Applied Neurobiology, 37(1), pp. 56–74. doi: 10.1111/j.1365-2990.2010.01139.x.

9 Bruhn, H., Fransson, P. and Frahm, J. (2001) ‘Modulation of cerebral blood oxygenation by indomethacin: MRI at rest and functional brain activation’, Journal of Magnetic Resonance Imaging, 13(3), pp. 325–334. doi: 10.1002/jmri.1047.

10 Buckner, R. L. et al. (1996) ‘Detection of cortical activation during averaged single trials of a cognitive task using functional magnetic resonance imaging’, Proceedings of the National Academy of Sciences, 93(25), pp. 14878–14883. doi: 10.1073/pnas.93.25.14878.

11 Buxton, R. B., Wong, E. C. and Frank, L. R. (1998) ‘Dynamics of Blood Flow and Oxygenation Changes During Brain Activation?: The Balloon Model’, (17), pp. 855–864.

12 Chen, B. R. et al. (2014) ‘A critical role for the vascular endothelium in functional neurovascular coupling in the brain’, Journal of the American Heart Association, 3(3), pp. 1–14. doi: 10.1161/JAHA.114.000787.

13 Chen, J. J. and Pike, G. B. (2009) ‘BOLD-specific cerebral blood volume and blood flow changes during neuronal activation in humans’, NMR in Biomedicine, 22(10), pp. 1054–1062. doi: 10.1002/nbm.1411.

14 Cohen, J. (2013) Statistical Power Analysis for the Behavioral Sciences. 2nd Editio. Routledge. doi: 10.4324/9780203771587.

15 D’Esposito, M. et al. (1999) ‘The effect of normal aging on the coupling of neural activity to the bold hemodynamic response’, NeuroImage, 10(1), pp. 6–14. doi: 10.1006/nimg.1999.0444.

16 D’Esposito, M., Deouell, L. Y. and Gazzaley, A. (2003) ‘Alterations in the BOLD fMRI signal with ageing and disease: A challenge for neuroimaging’, Nature Reviews Neuroscience, 4(11), pp. 863–872. doi: 10.1038/nrn1246.

17 Erdfelder, E. et al. (2009) ‘Statistical power analyses using G*Power 3.1: Tests for correlation and regression analyses’, Behavior Research Methods, 41(4), pp. 1149–1160. doi: 10.3758/BRM.41.4.1149.

18 Farkas, E. and Luiten, P. G. M. (2001) Cerebral microvascular pathology in aging and Alzheimer’s disease, Progress in Neurobiology. doi: 10.1016/S0301-0082(00)00068-X.

19 Félétou, M., Huang, Y. and Vanhoutte, P. M. (2011) ‘Endothelium-mediated control of vascular tone: COX-1 and COX-2 products’, British Journal of Pharmacology, 164(3), pp. 894–912. doi: 10.1111/j.1476-5381.2011.01276.x.

20 Friston, K. (2003) ‘Dynamic Causal Modelling’, in Human Brain Function: Second Edition, pp. 1063–1090. doi: 10.1016/B978-012264841-0/50054-8.

21 Friston, K. et al. (2007) ‘Variational free energy and the Laplace approximation’, NeuroImage, 34(1), pp. 220–234. doi: 10.1016/j.neuroimage.2006.08.035.

22 Friston, K. J. et al. (1995) ‘Analysis of fMRI Time-Series Revisited’, NeuroImage, 2(1), pp. 45–53. doi: 10.1006/nimg.1995.1007.

23 Friston, K. J. et al. (2000) ‘Nonlinear responses in fMRI: The balloon model, Volterra kernels, and other hemodynamics’, NeuroImage, 12(4), pp. 466–477. doi: 10.1006/nimg.2000.0630.

24 Geisser, S. and Greenhouse, S. W. (1959) ‘ON METHODS IN THE ANALYSIS OF PROFILE variance. Furthermore, an analysis of variance approach permits the analysis of a set of data which cannot be handled by multivariate procedures, namely, the case where n, the number of random vectors, is less t’, Psychometrika, 24(2).

25 Glover, G. H., Li, T.-Q. and Ress, D. (2000) ‘Image-based method for retrospective correction of physiological motion effects in fMRI: RETROICOR’, Magnetic Resonance in Medicine, 44(1), pp. 162–167. doi: 10.1002/1522-2594(200007)44:1<162::AID-MRM23>3.0.CO;2-E.

26 Handwerker, D. A. et al. (2012) ‘The continuing challenge of understanding and modeling hemodynamic variation in fMRI’, NeuroImage. Elsevier B.V., 62(2), pp. 1017–1023. doi: 10.1016/j.neuroimage.2012.02.015.

27 Handwerker, D. A., Ollinger, J. M. and D’Esposito, M. (2004) ‘Variation of BOLD hemodynamic responses across subjects and brain regions and their effects on statistical analyses’, NeuroImage, 21(4), pp. 1639–1651. doi: 10.1016/j.neuroimage.2003.11.029.

28 Havlicek, M. et al. (2015) ‘Physiologically informed dynamic causal modeling of fMRI data’, NeuroImage. Elsevier B.V., 122, pp. 355–372. doi: 10.1016/j.neuroimage.2015.07.078.

29 Haydon, P. G. and Carmignoto, G. (2006) ‘Astrocyte control of synaptic transmission and neurovascular coupling’, Physiological Reviews, 86(3), pp. 1009–1031. doi: 10.1152/physrev.00049.2005.

30 Heinzle, J. et al. (2016) ‘NeuroImage A hemodynamic model for layered BOLD signals’, NeuroImage. Elsevier Inc., 125, pp. 556–570. doi: 10.1016/j.neuroimage.2015.10.025.

31 Hesselmann, V. et al. (2001) ‘Age related signal decrease in functional magnetic resonance imaging during motor stimulation in humans’, Neuroscience Letters, 308(3), pp. 141–144. doi: 10.1016/S0304-3940(01)01920-6.

32 Hillman, E. M. C. (2014) ‘Coupling Mechanism and Significance of the BOLD Signal: A Status Report’, Annual Review of Neuroscience, 37(1), pp. 161–181. doi: 10.1146/annurev-neuro-071013-014111.

33 Huber, L. et al. (2014) ‘Investigation of the neurovascular coupling in positive and negative BOLD responses in human brain at 7T’, NeuroImage. Elsevier Inc., 97, pp. 349–362. doi: 10.1016/j.neuroimage.2014.04.022.

34 Huettel, S. A., Singerman, J. D. and McCarthy, G. (2001) ‘The effects of aging upon the hemodynamic response measured by functional MRI’, NeuroImage, 13(1), pp. 161–175. doi: 10.1006/nimg.2000.0675.

35 Kannurpatti, S. S. et al. (2010) ‘Neural and vascular variability and the fMRI-BOLD response in normal aging’, Magnetic Resonance Imaging. Elsevier Inc., 28(4), pp. 466–476. doi: 10.1016/j.mri.2009.12.007.

36 Kasper, L. et al. (2017) ‘The PhysIO Toolbox for Modeling Physiological Noise in fMRI Data’, Journal of Neuroscience Methods. Elsevier B.V., 276, pp. 56–72. doi: 10.1016/j.jneumeth.2016.10.019.

37 Kiebel, S. J. et al. (2007) ‘Dynamic causal modeling: A generative model of slice timing in fMRI’, NeuroImage, 34(4), pp. 1487–1496. doi: 10.1016/j.neuroimage.2006.10.026.

38 Kim, S. G. and Ogawa, S. (2012) ‘Biophysical and physiological origins of blood oxygenation level-dependent fMRI signals’, Journal of Cerebral Blood Flow and Metabolism. Nature Publishing Group, 32(7), pp. 1188–1206. doi: 10.1038/jcbfm.2012.23.

39 Kröger, I. L. and May, A. (2014) ‘Central effects of acetylsalicylic acid on trigeminal-nociceptive stimuli’, pp. 1–6. doi: 10.1186/1129-2377-15-59.

40 Lauritzen, M. (2005) ‘Reading vascular changes in brain imaging: Is dendritic calcium the key?’, Nature Reviews Neuroscience, 6(1), pp. 77–85. doi: 10.1038/nrn1589.

41 Lind, B. L. et al. (2013) ‘Rapid stimulus-evoked astrocyte Ca2+ elevations and hemodynamic responses in mouse somatosensory cortex in vivo’, Proceedings of the National Academy of Sciences, 110(48), pp. E4678–E4687. doi: 10.1073/pnas.1310065110.

42 Maihöfner, C. et al. (2007) ‘Brain imaging of analgesic and antihyperalgesic effects of cyclooxygenase inhibition in an experimental human pain model: A functional MRI study’, European Journal of Neuroscience, 26(5), pp. 1344–1356. doi: 10.1111/j.1460-9568.2007.05733.x.

43 Maldjian, J. A. et al. (2003) ‘An automated method for neuroanatomic and cytoarchitectonic atlas-based interrogation of fMRI data sets’, 19, pp. 1233–1239. doi: 10.1016/S1053-8119(03)00169-1.

44 Mandeville, J. B. et al. (1998) ‘Dynamic functional imaging of relative CBV during rat fore paw stimulation.’, Magn Res Med, 39(24), pp. 619–624.

45 Markus, H. S., Vallance, P. and Brown, M. M. (1994) ‘Differential effect of three cyclooxygenase inhibitors on human cerebral blood flow velocity and carbon dioxide reactivity’, Stroke, 25(9), pp. 1760–1764. doi: 10.1161/01.STR.25.9.1760.

46 Mauchly, J. W. (1940) ‘Significance Test for Sphericity of a Normal n-Variate Distribution’, The Annals of Mathematical Statistics, 11(2), pp. 204–209.

47 Meier-Ruge, W. et al. (1980) ‘Stereological changes in the capillary network and nerve cells of the aging human brain’, Mechanisms of Ageing and Development, 14(1–2), pp. 233–243. doi: 10.1016/0047-6374(80)90123-2.

48 Miller, K. L. et al. (2001) ‘Nonlinear temporal dynamics of the cerebral blood flow response’, Human Brain Mapping, 13(1), pp. 1–12. doi: 10.1002/hbm.1020.

49 Oláh, O. et al. (2012) ‘Regional differences in the neuronal expression of cyclooxygenase-2 (COX-2) in the newborn pig brain’, Acta Histochemica et Cytochemica, 45(3), pp. 187–192. doi: 10.1267/ahc.11056.

50 Penny, W. D. et al. (2010) ‘Comparing Families of Dynamic Causal Models’, 6(3). doi: 10.1371/journal.pcbi.1000709.

51 Quintana, A. et al. (1983) ‘Effects of aspirin and indomethacin on cerebral circulation in the conscious rat: Evidence for a physiological role of endogenous prostaglandins’, Prostaglandins, 25(4), pp. 549–556. doi: 10.1016/0090-6980(83)90027-8.

52 Riera, J. J. and Sumiyoshi, A. (2010) ‘Brain oscillations: Ideal scenery to understand the neurovascular coupling’, Current Opinion in Neurology, 23(4), pp. 374–381. doi: 10.1097/WCO.0b013e32833b769f.

53 Stefanovic, B., Bosetti, F. and Silva, A. C. (2006) ‘Modulatory role of cyclooxygenase-2 in cerebrovascular coupling’, NeuroImage, 32(1), pp. 23–32. doi: 10.1016/j.neuroimage.2006.03.014.

54 Stephan, K. E. et al. (2007) ‘Comparing hemodynamic models with DCM’, NeuroImage. Elsevier Inc., 38(3), pp. 387–401. doi: 10.1016/j.neuroimage.2007.07.040.

55 Sudlow, C. et al. (2015) ‘UK Biobank?: An Open Access Resource for Identifying the Causes of a Wide Range of Complex Diseases of Middle and Old Age’, pp. 1–10. doi: 10.1371/journal.pmed.1001779.

56 Tallini, Y. N. et al. (2007) ‘Propagated endothelial Ca2+ waves and arteriolar dilation in vivo: Measurements in Cx40BAC-GCaMP2 transgenic mice’, Circulation Research, 101(12), pp. 1300–1309. doi: 10.1161/CIRCRESAHA.107.149484.

57 Tsubokura, S. et al. (1991) ‘Localization of prostaglandin endoperoxide synthase in neurons and glia in monkey brain’, Brain Research, 543(1), pp. 15–24. doi: 10.1016/0006-8993(91)91043-Z.

58 Vane, J. R. (1971) ‘Inhibition of Prostaglandin Synthesis as a Mechanism of Action for Aspirin-like Drugs’, Nature New Biology, 231(25), pp. 232–235. doi: 10.1038/newbio231232a0.

59 Vane, J. R. and Botting, R. M. (2003) ‘The mechanism of action of aspirin’, 110, pp. 255–258. doi: 10.1016/S0049-3848(03)00379-7.

60 Vanhoutte, P. M. (2009) ‘COX-1 and vascular disease’, Clinical Pharmacology and Therapeutics. Nature Publishing Group, 86(2), pp. 212–215. doi: 10.1038/clpt.2009.108.

61 De Vries, E. F. J. et al. (2003) ‘Synthesis and in vivo evaluation of 18F-desbromo-dup-697 as a PET tracer for cyclooxygenase-2 expression’, Journal of Nuclear Medicine, 44(10), pp. 1700–1706.

62 Wang, X. et al. (2006) ‘Astrocytic Ca 2+ signaling evoked by sensory stimulation in vivo’, Nature Neuroscience, 9(6), pp. 816–823. doi: 10.1038/nn1703.

63 Warner, T. D., Nylander, S. and Whatling, C. (2011) ‘Anti-platelet therapy: Cyclo-oxygenase inhibition and the use of aspirin with particular regard to dual anti-platelet therapy’, British Journal of Clinical Pharmacology, 72(4), pp. 619–633. doi: 10.1111/j.1365-2125.2011.03943.x.

64 West, K. L. et al. (2019) ‘NeuroImage BOLD hemodynamic response function changes significantly with healthy aging’, NeuroImage. Elsevier Inc., 188(September 2018), pp. 198–207. doi: 10.1016/j.neuroimage.2018.12.012.

65 Winship, I. R., Plaa, N. and Murphy, T. H. (2007) ‘Rapid Astrocyte Calcium Signals Correlate with Neuronal Activity and Onset of the Hemodynamic Response In Vivo’, Journal of Neuroscience, 27(23), pp. 6268–6272. doi: 10.1523/jneurosci.4801-06.2007.

66 Wölfle, S. E. et al. (2011) ‘Non-linear relationship between hyperpolarisation and relaxation enables long distance propagation of vasodilatation’, Journal of Physiology, 589(10), pp. 2607–2623. doi: 10.1113/jphysiol.2010.202580.

67 Wright, M. E., Wise, R. G. and Harris, J. J. (2018) ‘Can Blood Oxygenation Level Dependent Functional Magnetic Resonance Imaging Be Used Accurately to Compare Older and Younger Populations? A Mini Literature Review’, 10(November), pp. 1–7. doi: 10.3389/fnagi.2018.00371.

68 Yasojima, K. et al. (1999) ‘Distribution of cyclooxygenase-1 and cyclooxygenase-2 mRNAs and proteins in human brain and peripheral organs’, Brain Research, 830(2), pp. 226–236. doi: 10.1016/S0006-8993(99)01389-X.

69 Yermakova, A. V. et al. (1999) ‘Cyclooxygenase-1 in Human Alzheimer and Control Brain: Quantitative Analysis of Expression by Microglia and CA3 Hippocampal Neurons’, Journal of Neuropathology and Experimental Neurology, 58(11), pp. 1135–1146. doi: 10.1097/00005072-199911000-00003.

70 Zirpel, L., Lachica, E. and Rubel, E. (1995) ‘Activation of a metabotropic glutamate receptor increases intracellular calcium concentrations in neurons of the avian cochlear nucleus’, The Journal of Neuroscience, 15(1), pp. 214–222. doi: 10.1523/JNEUROSCI.15-01-00214.1995.

